# Immunogenicity and safety of Biological E’s CORBEVAX™ vaccine as a heterologous booster dose in adult volunteers previously vaccinated with two doses of either COVISHIELD™ or COVAXIN: A Prospective double-blind randomised phase III clinical study

**DOI:** 10.1101/2022.12.29.22284049

**Authors:** Subhash Thuluva, Vikram Paradkar, SubbaReddy Gunneri, Vijay Yerroju, Rammohan Mogulla, Pothakamuri Venkata Suneetha, Kishore Turaga, Akshay Binayke, Aymaan Zaheer, Amit Awasthi, Rashmi Virkar, Manish Narang, Pradeep Nanjappa, Niranjana Mahantshetti, BishanSwarup Garg, Mandal RavindraNath Ravi

**Author notes:** **Corresponding Author Name:** Subhash Thuluva, Biological E Limited, 18/1&3, Azamabad, Hyderabad – 500 020, Telangana, India. **E-mail**. **Data Sharing Agreement** Additional study data which is not part of the manuscript can be made available upon request and addressed to the corresponding author Dr. Subhash Thuluva at his. **Authors’ Contribution:** ST and VP conceptualized the study and edited the manuscript for intellectual content. ST, SG, VY, RM, PVS and KT curated, accessed and verified the data and helped in interim report generation. VP led the immunogenicity experiments. RV coordinated the execution of neutralizing antibody assays. AB, AZ and AA contributed in performing the ELISpot testing for cellular immune response assessment. MS, PN, NM, BSG and MRNR were the key contributors of study conduct. ST was responsible for overall supervision of the project. All authors contributed to data interpretation, reviewing, and editing this manuscript.

## Abstract

**Background:** Vaccines developed against Covid-19 infection were effective in controlling symptomatic infections and hospitalizations. However, waning immunity was reported within 6 months of primary vaccination series. Due to waning of SARS-CoV-2 specific primary immunity, protection towards emerging variants of concern (VoC) was low. To rejuvenate the immunogenicity of vaccines, a third or booster dose was highly recommended by many state governments. In this regard, several clinical studies were conducted to evaluate the homologous or heterologous booster dose effectiveness against VoCs and showed that heterologous immune boosting more effective in controlling breakthrough infections. In this study, we studied the safety and immunogenicity of Biological-E’s CORBEVAX™ vaccine in adult population as a heterologous booster dose.

**Methods:** This is a prospective phase-3, randomised, double-blind, placebo-controlled, study evaluating safety, reactogenicity, tolerability and immunogenicity of CORBEVAX™ vaccine as a heterologous booster dose administered to adult volunteers previously vaccinated with two doses of either COVISHIELD™ or COVAXIN at least 6 months ago. Subjects were RT-PCR negative to SARS-CoV-2 prior to enrolment. A total of 416 subjects between 18 to 80 years of age, were enrolled in to one of the two treatment (COVISHIELD™ or COVAXIN primed subjects) groups (n=208/group) for safety and immunogenicity assessment. Within each group (n=208), subjects were randomized to receive CORBEVAX™ vaccine or placebo in a 3:1 ratio.

**Findings:** The safety profile of CORBEVAX™ vaccine administered as booster dose is comparable to the placebo-control group. All the reported adverse events (AEs) were mild to moderate in their intensity. There was no grade 3 or serious or AEs of special interest (AESI) reported during the study period and all the reported AEs resolved without any sequelae. CORBEVAX™ booster dose administration resulted in significant increase in humoral immune response (nAb titers and Anti-RBD IgG concentration) that was much superior to the placebo in both COVISHIELD™ and COVAXIN recipient arms. Significant increase in nAb titers against Omicron VOC as well as cellular immune response was also observed post CORBEVAX™ booster dose administration.

**Interpretations:** Enhancement of immune response coupled with excellent safety profile of the CORBEVAX™ booster dose demonstrates significant benefit of giving CORBEVAX™ heterologous booster dose to subjects that have received COVISHIELD™ or COVAXIN primary vaccination; as early as 6 months post second dose of primary vaccination.

The study was prospectively registered with clinical trial registry of India-CTRI/2022/01/039366

## INTRODUCTION

Spike glycoprotein of SARS-CoV-2 ancestral strain is the main candidate protein that is used as a target for vaccine development. Antibodies directed against the spike protein were shown to provide high rate of protection against developing symptomatic infection and hospitalizations caused due to SARS-CoV-2 strain^1^. Various SARS-CoV-2 variants have emerged worldwide with mutations in the spike protein. Increased number of breakthrough infections with Delta and Omicron variants of SRS-CoV-2 in fully vaccinated individuals was observed. Breakthrough infections were associated with immune escape and also waning of primary immune responses elicited by vaccines or natural infection. Six months after receiving two doses of BNT162b2 vaccine, vaccine effectiveness (VE) against ancestral strain was 66% where as VE against Omicron variant was only 9%. Similar reduction in VE was also noticed with mRNA-1273 and ChAdOx1. No protective effect against symptomatic disease caused by the omicron variant was noted after six months from administration of the second dose of ChAdOx1^2^. To resolve the ongoing pandemic by augmenting the waning immune responses, periodic third or booster dose administration was recommended by many countries. Booster vaccines enhance the immunity and expand the magnitude and breadth of immune responses against variants of concern^3^.

Neutralizing antibody titers against wild-type SARS-CoV-2 virus were increased by a factor of 5.5 and 3.8, respectively after homologous BNT162b2 and mRNA-1273 booster dose vaccination, respectively, given six months after the second dose administration^4,5^. Third dose of BNT162b2 vaccine was highly effective in preventing hospitalization and deaths^6^. This phenomenon was also supported by another observational study in elderly population from Israel. Authors have shown that a booster dose reduced the rate of confirmed infections and severe disease by a factor of 11.3 and 19.5, respectively^7^. Data from different clinical trials conducted globally indicates that heterologous boosting increased the levels of binding and neutralizing antibodies and proven to be beneficial in inducing durable and protective immunity compared to the homologous booster dose. A study by Atmar et al, has shown that homologous boosters increased neutralizing antibody titers by a factor of 4 to 20, whereas heterologous boosters increased titers by a factor of 6 to 73^8^. In a study involving 4,806,026 veterans testing real world vaccine effectiveness of homologous and heterologous booster dose, large number of breakthrough infections were documented in participants who had received a homologous Ad26.COV2.S booster and the infection rate was lower in participants who were boosted with a heterologous mRNA vaccine^9^. The UK COMCOV trial demonstrated that a heterologous schedule can be more immunogenic than a homologous schedule (ChAd/BNT with ChAd/ChAd, and BNT/ChAd with BNT/BNT)^10^.

In India, >90% of adult population received either COVAXIN or Covishiled as a primary vaccination and a precautionary third dose was recommended by the Indian government for elderly population (Jan, 2022). However, there is limited data available on safety and immunogenicity of a third or booster dose in India. In the current phase-3 clinical study, safety and immunogenicity of a protein sub-unit vaccine (CORBEVAX™) developed by Biological E limited was tested when administered as a heterologous booster dose in subjects who received either COVAXIN or COVISHIELD™ as a primary vaccination series. Safety and immunogenicity of CORBEVAX™ vaccine administered as primary vaccination in healthy individuals aged 5-80 years was well established in different phase 1-3 studies and shown to be well tolerated, safe and can induce SARS-CoV-2 specific humoral and cellular immune responses with estimated vaccine effectiveness of >90% in preventing symptomatic COVID-19 infection^11,12.^

## METHODS

### Study Design and Study Population

A prospective, multicenter, placebo controlled, double-blind, randomized phase III clinical study was conducted to evaluate the immunogenicity and safety of single heterologous booster dose of Biological E’s CORBEVAX™ vaccine.The protocol was approved by the investigational review board or ethics committee at each study site and the study was conducted in seven sites across India in accordance with the principles defined in the Declaration of Helsinki, International Conference on Harmonization guidelines (Good Clinical Practices), and the local regulatory guidelines.

Adults aged between 18 to 80 years who were negative to SARS-CoV-2 infection at the time of enrolment and received primary vaccination series with two doses of either inactivated vaccine (COVAXIN: Group-1) or viral vector based vaccine (COVISHIELD™: Group-2) at least 6 months (+28 days) ago were eligible for participation. In total, 546 subjects were screened to enroll 416 subjects into one of the two treatment groups (n=208/group). In each group, 156 subjects received heterologous single booster dose of CORBEVAX™ vaccine and 52 subjects received placebo in a 3:1 ratio. Subjects with high risk or comorbid condition and healthy individuals were equally enrolled into the study. All participants provided written informed consent before enrollment into the study. There were no major protocol deviations reported at any of the study sites during the conduct of the study. The average study duration was 28 days (+4 days) for each subject and were scheduled to be followed up for 6 months and 9 months after vaccination.

### Blinding and Randomization

Double blinding was paired to randomization, where participants were randomly assigned to one of the two vaccination arms (CORBEVAX™ or placebo) according to a random generated algorithm. Subjects were recruited by vaccination unit, age group and absence/presence of comorbidities. Participants and investigators were blinded to group allocation. The test (CORBEVAX™) and placebo were labelled and packed identical to each other.The interactive web response system (IWRS) assigned a unique intervention code, which dictated the intervention assignment for the participant. Except independent statistician from contract research organization (CRO) and personnel responsible for conducting packaging/labelling/blinding of the investigational product, all the personnel involved in the study remained blinded to the study vaccine.

### Procedure

Biological E’s CORBEVAX™ vaccine is a recombinant sub unit vaccine consists of RBD protein as an antigen, CpG-1018 and Aluminium hydroxide as adjuvants ^11^. A 0.5mL of CORBEVAX™ or placebo (adjuvant formulated in Tris-buffer) was administered via an intramuscular (IM) injection into the deltoid muscle. Prophylactic medication is not prescribed either before or after vaccination.

### Outcomes

The primary objective of the study was to assess the immunogenicity in terms of virus neutralizing antibodies (VNA) 28 days after single booster dose of CORBEVAX™ in comparison with placebo. Secondary objective was to assess safety, tolerability and reactogenicity, after single booster dose of CORBEVAX™ for first 28 days and for further up to 6 months.

Other secondary outcomes were to assess: immunogenic superiority of CORBEVAX™ against Placebo in terms of virus neutralizing antibodies (VNA); geometric mean titre (GMT/C), geometric mean fold rise (GMFR) of anti-RBD IgG against SARS-CoV-2; immune response against Delta variant at28 days’ post booster dose. Assessing immune responses against beta variant and cellular immune responses at 28 days after booster dose were the exploratory objectives of the study.

### Safety assessments

The secondary safety endpoints were solicited local reactions at the injection site, solicited systemic reactions and the proportion of subjects with at least one unsolicited treatment emergent adverse event (TEAE), serious adverse event (SAE) and medically attended adverse event (MAAE)and adverse events of special interest (AESI) reported in the study. Occurrence, severity (mild, moderate, severe and life threatening) and relatedness of all adverse reactions were collected. All participants were observed for at least 60 minutesfor any immediate safety concerns at study center. Solicited local and general AEs were recorded for 7 consecutive days (Day 0-6), captured through subject diary after booster dose. Unsolicited AEs, if any were collected during the first 28 days after the booster dose and further up to 6 months follow up period.

### Immunogenicity Analysis

Humoral immune responses were assessed by measuring anti-RBD IgG levels and neutralizing antibody (nAb) titers. Subject sera samples collected prior to vaccination (Day-0) and 28 days after booster dose administration (CORBEVAX™ or Placebo) were tested in an ELISA assay to determine the concentration of antibodies generated against the RBD antigen. For each subject, Anti-RBD IgG concentrations were recorded in ELISA Units/mL. From this data, fold rise in antibody concentration and geometric mean concentrations (GMC’s) were calculated.

Neutralizing Antibody (nAb) titers against the Ancestral SARS-CoV-2 (Wuhan-D614G) strainwas assessed by the Plaque Reduction Neutralization Titer (PRNT) method tested at IRSHA Laboratories, Pune, India via a validated assay. For each sera sample, the PRNT_50_ titer was determined i.e. the dilution of the sera that can neutralize 50% of virus infectivity. Subject sera samples collected prior to vaccination (Day-0) and 28 days after booster dose administration were tested in the assay to determine the nAb titers. From this data, fold rise in nAb titer was determined. Then, geometric mean titers (GMT’s) were calculated at Day-0 and Day-28 time-points for subjects in both COVISHIELD™ and COVAXIN groups after the booster dose with CORBEVAX™ or Placebo.

Neutralizing Antibody (nAb) titers against the Omicron variant of concern was assessed by the Pseudovirus Neutralization Assay (PNA) method tested at Dr Reddy’s Institute of Life Sciences, Hyderabad, India. A HIV Lentivirus-based pseudovirus expressing Omicron (BA.1.159 or BA.1) Spike protein was used for neutralization studies. For each sera sample, the PNA_50_ titer was determined at Day-0 and Day-28 after booster dose administration. From this data, fold rise in nAb titer and GMT’s were calculated for Day-0 and Day-28 time-points. Subject sera samples with detectable level of PNA titer were categorized as responders and % of subjects that demonstrated detectable PNA titers was calculated at Day-0 and Day-28 time-points. Due to limited testing capacity, only subjects that were dosed with CORBEVAX™ in both groups were tested and hence statistical comparison can’t be conducted.

Cellular immune responses were measured by stimulating isolated peripheral blood mononuclear cells (PBMCs) with different antigens viz. SARS-CoV-2 RBD peptides (JPT, Berlin, Germany) as specific antigen; DMSO as non-specific stimulant and PHA protein as common stimulant for assay validity. After stimulation, the number of PBMC’s secreting Interferon-gamma cytokine were detected in ELISPOT and the PBMC’s responding specifically to SARS-CoV-2 RBD peptides were quantified after subtracting the non-specific contribution from the DMSO response. The total Interferon-gamma reactive PBMC’s were quantified as Spot Forming Units (SFU’s) per million PBMC’sby enzyme linked immunosorbent spot (ELISPOT) and enzyme linked immunosorbent assay (ELISA). Apart from ELISpot assay, cellular immune responses were assessed by measuring cytokine secretion using TrueCulture tubes^13^. Whole blood samples were incubated in TrueCulture tubes coated with SARS-COV-2 peptides (Myriad Inc., TX, USA). The cytokines secreted by the PBMC’s present in the blood samples were then measured by ELISA method using kits supplied by Becton Dickinson Diagnostics.

### Statistical Analyses

#### Sample Size Determination

The sample size was driven by the primary immunogenicity endpoint evaluation. There were two treatment groups viz., Group-1 (n=208) (COVISHIELD™ primed) and Group-2 (n=208) (COVAXIN primed). Subjects within each of these two treatment groups was further stratified in 3:1 ratio, either to receive the CORBEVAX™ booster dose (n=156) or a placebo (n=52). A proportion (% ≥2-fold increase) was measured in each group under both CORBEVAX™ and placebo treatment arms. A total of two hypothesis tests were anticipated, each comparing a treatment arm with its corresponding placebo arm using a simple z-test of the difference between two proportions. Two tests were performed viz., treatment arm versus placebo arm under each group. The type I error rate was controlled using a Bonferroni adjustment. It is assumed that the proportion of subjects achieving a ≥2-fold increase in virus neutralization assay in each of the two groups within each treatment arm would be 80% respectively for power computation.

A one-way study was planned to test whether each of the two treatment sub-group proportions were significantly different from their corresponding placebo-group proportions at the 2.5% significance level. The total sample size before dropout allocation was calculated to be 336 subjects with 42 subjects and 42 subjects required in each of the two placebo sub-groups and 126 and 126 subjects in each of the two treatment sub-groups respectively. The corresponding powers of each of the tests for comparison are 0.901 and 0.901 respectively. A 15% dropout rate is assumed, which is the percentage of subjects that are expected to be lost at random during the course of the study. Accordingly, the total sample size would then be 416 subjects, with 156 each in the two treatment sub-groups and 52 each in their corresponding two placebo sub-groups respectively.

The descriptive analysis of demographics and other baseline characteristics was performed on Intent-to-treat (ITT) population (participants who were randomized). All safety-related tabulations and analyses were performed on safety population (subjects who received at least one dose of any study vaccine). Continuous variables were summarized by group using the descriptive statistics: number of subjects overall and with missing and non-missing data (n), mean, standard deviation (SD), median, minimum (min) and maximum (max). Categorical variables were summarized by group presenting absolute and relative frequencies (n and %) of subjects in each category. Percentages were presented with exact 95% Clopper-Pearson confidence intervals (CIs). For the confidence interval estimation of proportions, the exact binomial calculations were applied. Safety data were summarized by System Organ Class and Preferred Term. Serious adverse events, related adverse events, adverse events leading to death or withdrawal, solicited adverse events, MAAEs and AESI were summarized separately.

The primary immunogenicity analysis was performed in the per-protocol population (participants who received the single booster dose and for whom protocol specified immunogenicity data was available). The geometric mean concentration/titer (GMC/T) was calculated as the mean of the logarithm of the neutralizing or IgG antibodies, back-transformed into the original scale. Geometric mean fold rise (GMFR) in anti-SARS-CoV IgG antibody titers and virus neutralizing antibodies (VNA) at day 28 from pre-booster, along with their corresponding 2-sided 95% CIs, was presented. Geometric mean fold rise (GMFR) is calculated as the mean of the difference of logarithmically transformed assay results (post vaccination time point – pre vaccination time point) and back transformed into the original scale.

Proportion of subjects achieving ≥2-fold rise in virus neutralizing antibodies (VNA) and anti SARS-CoV IgG antibody titres at 28 days from pre-booster levels, along with their corresponding 2-sided 95% CIs, was presented for all the treatment groups. Seroconversion (SP) was defined as proportion of subjects with ≥2-fold rise in virus neutralising antibodies (VNA) from pre-booster levels at 28 days after single booster dose. Immunogenic superiority of CORBEVAX™ against placebo (secondary end point) was defined as the lower limit of the two-sided 95% confidence interval for the ratio (Test/Reference) of the two geometric mean concentrations or titers above 1. All the statistical analysis was conducted by the sponsor approved CRO using SAS® 9.4 or higher.

## RESULTS

### Participants

The subjects’ demographics and baseline characteristics are described in Table 1, and their disposition is shown in Figure 1. Study was conducted between January 2022 and August 2022.

**TABLE 1:**
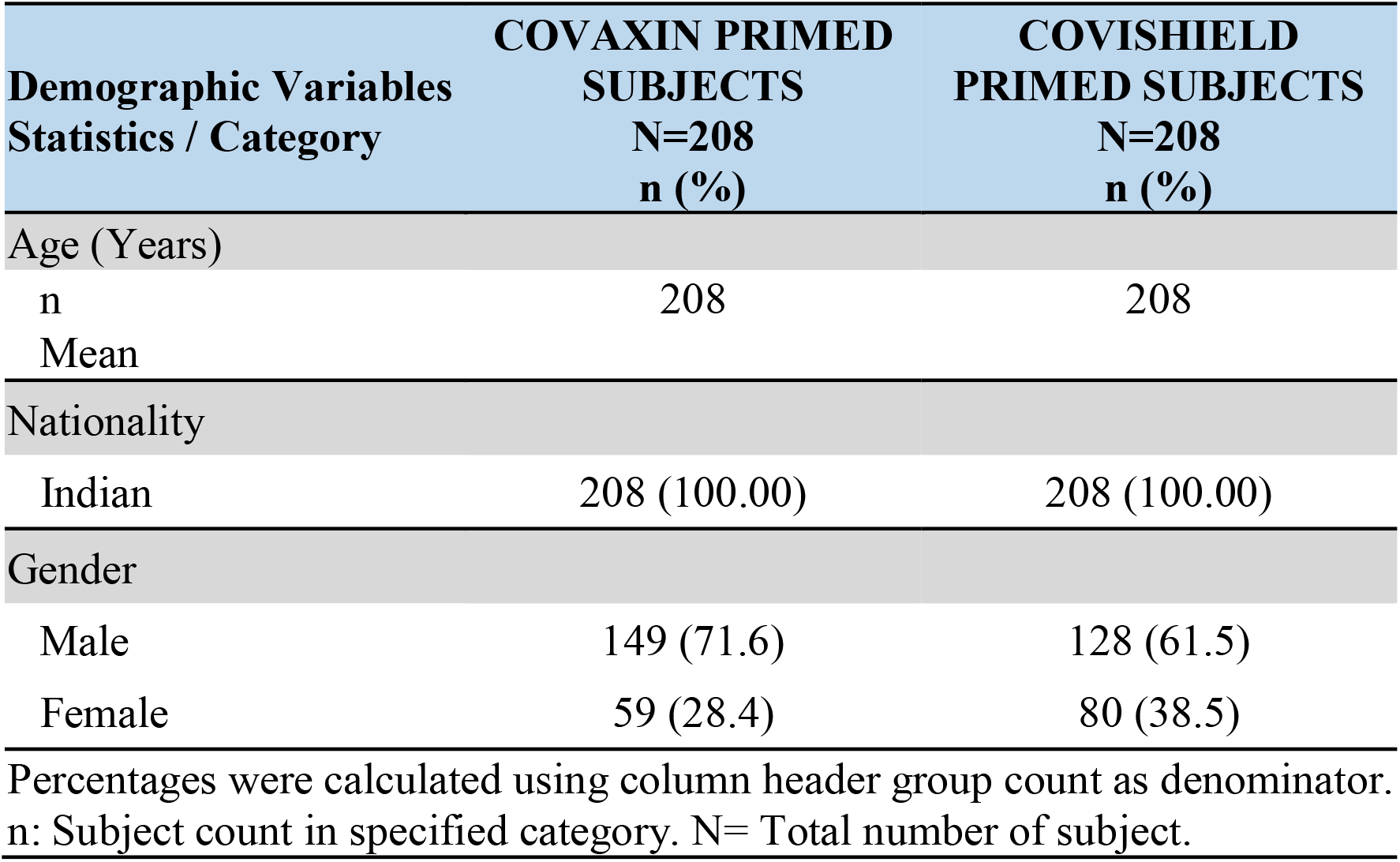
Demographic characteristics of study participants.

**Fig 1:**
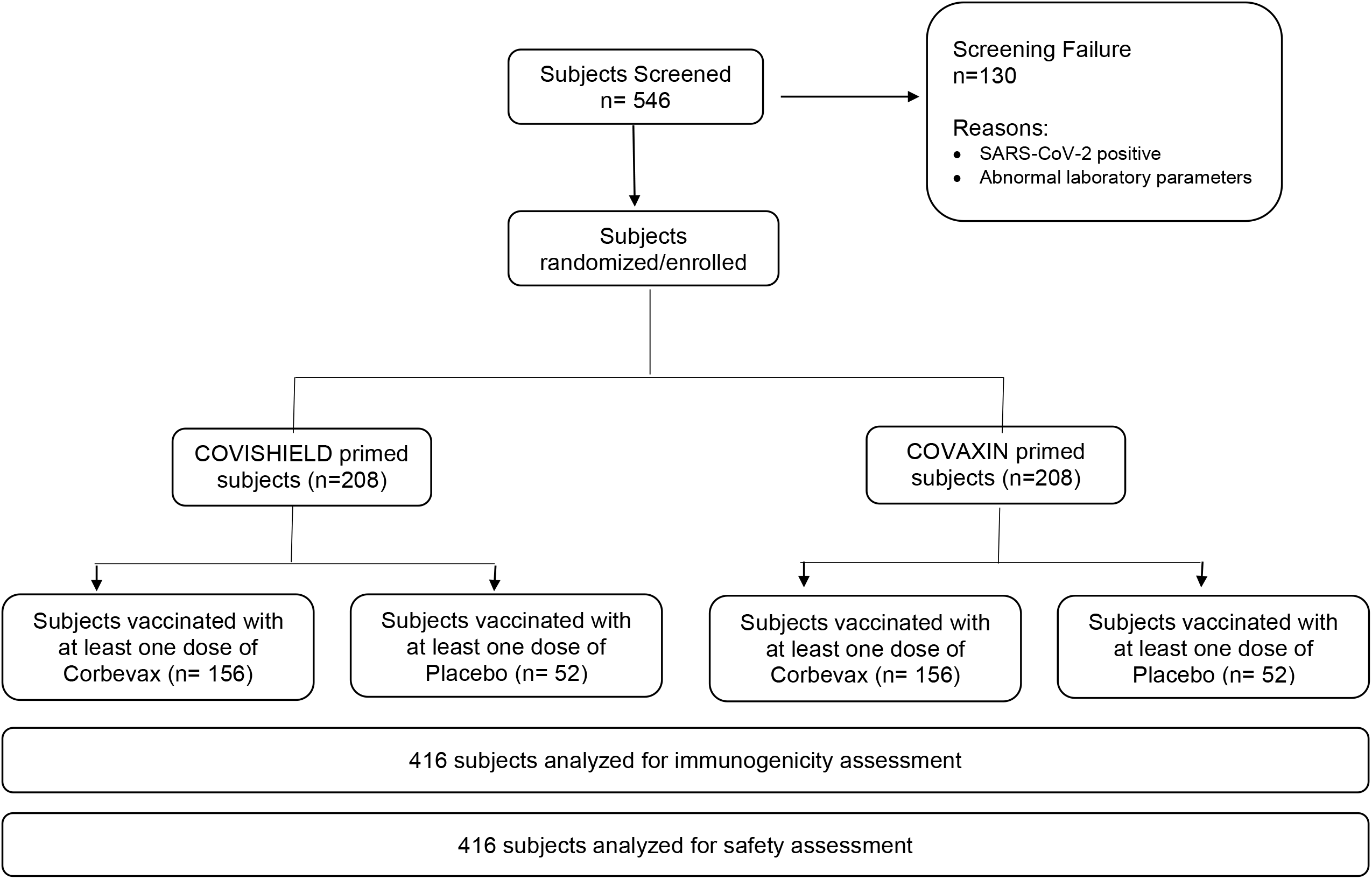
Study subjects disposition.

### Safety Findings

In COVAXIN primed subjects, 32 (20.5%) out of 156 subjects and 7 (13.5%) out of 52 subjects reported at least one adverse event after booster dose with CORBEVAX™ or Placebo respectively. In COVISHIELD™ primed subjects, 28 (17.9%) out of 156 and 8 (15.4%) out of 52 subjects reported at least one adverse event after booster dose with CORBEVAX™ or Placebo respectively. Overview of the subjects with AEs in total vaccinated group (TVC) is presented in table 2. Summary of AEs by system organ class (SOC) and preferred term (PT) in subjects vaccinated with CORBEVAX™ vaccine or placebo in COVAXIN and COVISHIELD™ primed subjects is presented in table 3. All the reported AEs were mild to moderate in their intensity. There were no serious AEs/grade-3 AEs/deaths/AESIs reported during the study period and all the reported AEs resolved without any sequelae. Severity and causality of reported AEs were presented in table 4. Most of the AEs were related to vaccination. None of the subjects discontinued the study due to an adverse event. Overall, safety profile of CORBEVAX™ vaccine as a heterologous booster dose found to be safe and well tolerated in adults who were primed with COVISHIELD™ or COVAXIN vaccines.

**Table 2:** Overview of subjects with AEs – Total Vaccinated Group.

| Category | COVAXIN PRIMED<br>SUBJECTS |  | COVISHIELD PRIMED<br>SUBJECTS |  |
| --- | --- | --- | --- | --- |
|  | CORBEVAX<br>N=156 n (%) | PLACEBO<br>N=52 n (%) | CORBEVAX<br>N=156 n (%) | PLACEBO<br>N=52 n (%) |
| Overall AEs | 32 (20.5) 44 | 7 (13.5) 10 | 28 (17.9) 38 | 8 (15.4) 8 |
| Local AEs | 16 (10.3) 17 | 3 (5.8) 3 | 19 (12.2) 21 | 7 (13.5) 7 |
| Systemic AEs | 23 (14.7) 27 | 6 (11.5) 7 | 13 (8.3) 17 | 1 (1.9) 1 |
| Any Medically<br>Attended AEs | 0 (0.0) 0 | 0 (0.0) 0 | 0 (0.0) 0 | 0 (0.0) 0 |
| AEs within 60 minutes<br>postvac | 0 (0.0) 0 | 0 (0.0) 0 | 1 (0.6) 1 | 2 (3.8) 2 |
| AEs within 7 days<br>postvac | 32 (20.5) 44 | 7 (13.5) 10 | 28 (17.9) 38 | 8 (15.4) 8 |
| Solicited AEs | 32 (20.5) 43 | 7 (13.5) 10 | 28 (17.9) 38 | 8 (15.4) 8 |
| Unsolicited AEs | 1 (0.6%) 1 | 0 (0.0) 0 | 0 (0.0) 0 | 0 (0.0) 0 |

**Table 3:** Summary of AEs by SOC and PT in subjects vaccinated with CORBEVAX™ vaccine or Placebo in Covaxin and Covishield primed subjects.

| SOC/PT<br>N<sub>1</sub> (%) n | COVAXIN PRIMED<br>SUBJECTS |  | COVISHIELD PRIMED<br>SUBJECTS |  |
| --- | --- | --- | --- | --- |
|  | CORBEVAX<br>N=156 n (%) | PLACEBO<br>N=52 n (%) | CORBEVAX<br>N=156 n (%) | PLACEBO<br>N=52 n (%) |
| OVERALL | 32 (20.5) 44 | 7 (13.5) 10 | 28 (17.9) 38 | 8 (15.4) 8 |
| Gastrointestinal disorders | 1 (0.6) 1 | 0 (0.0) 0 | 0 (0.0) 0 | 0 (0.0) 0 |
| Dyspepsia | 1 (0.6) 1 | 0 (0.0) 0 | 0 (0.0) 0 | 0 (0.0) 0 |
| General disorders and<br>administration site conditions | 27 (17.3) 29 | 3 (5.8) 5 | 23 (14.7) 28 | 8 (15.4) 8 |
| Chills | 1 (0.6) 1 | 1 (1.9) 1 | 0 (0.0) 0 | 0 (0.0) 0 |
| Injection site erythema | 2 (1.3) 2 | 0 (0.0) 0 | 2 (1.3) 2 | 1 (1.9) 1 |
| Injection site pain | 14 (8.9) 14 | 3 (5.8) 3 | 16 (10.3) 16 | 5 (9.6) 5 |
| Injection site swelling | 1 (0.6) 1 | 0 (0.0) 0 | 2 (1.3) 2 | 0 (0.0) 0 |
| Injection site itching | 0 (0.0) 0 | 0 (0.0) 0 | 1 (0.6) 1 | 1 (1.9) 1 |
| Pyrexia | 11 (7.1) 11 | 1 (1.9) 1 | 2 (1.3) 2 | 1 (1.9) 1 |
| Irritability post vaccinal | 0 (0.0) 0 | 0 (0.0) 0 | 2 (1.3) 2 | 0 (0.0) 0 |
| Fatigue | 0 (0.0) 0 | 0 (0.0) 0 | 3 (1.9) 3 | 0 (0.0) 0 |
| Musculoskeletal and connective<br>tissue disorders | 3 (1.92) 3 | 0 (0.0) 0 | 4 (2.6) 5 | 0 (0.0) 0 |
| Arthralgia | 3 (1.92) 3 | 0 (0.0) 0 | 2 (1.3) 2 | 0 (0.0) 0 |
| Myalgia | 0 (0.0) 0 | 0 (0.0) 0 | 3 (1.9) 3 | 0 (0.0) 0 |
| Respiratory, thoracic and<br>mediastinal disorders | 3 (1.92) 3 | 0 (0.0) 0 | 1 (0.6) 1 | 0 (0.0) 0 |
| Rhinorrhoea | 3 (1.92) 3 | 0 (0.0) 0 | 1 (0.6) 1 | 0 (0.0) 0 |
| Nervous system disorders | 8 (3.6) 8 | 5 (9.6) 5 | 3 (1.9) 3 | 0 (0.0) 0 |
| Headache | 8 (3.6) 8 | 5 (9.6) 5 | 3 (1.9) 3 | 0 (0.0) 0 |
| Skin and subcutaneous tissue<br>disorders | 0 (0.0) 0 | 0 (0.0) 0 | 1 (0.6) 1 | 0 (0.0) 0 |
| Pruritus | 0 (0.0) 0 | 0 (0.0) 0 | 1 (0.6) 1 | 0 (0.0) 0 |

**Table 4:** Overview of AEs by Severity & Causality.

| Category | COVAXIN<br>PRIMED<br>SUBJECTS |  | COVISHIELD<br>PRIMED<br>SUBJECTS |  |
| --- | --- | --- | --- | --- |
|  | CORBEV | PLACEB | CORBEV | PLACEB |
|  | AX<br>N=156 n<br>(%) | O<br>N=52 n<br>(%) | AX<br>N=156 n<br>(%) | O<br>N=52 n<br>(%) |
| Number of subjects with at least one AE | 32 (20.5)<br>44 | 7 (13.5)<br>10 | 28 (17.9)<br>38 | 8 (15.4)<br>8 |
| Number of subjects with at least one SAE | 0 (0.0) 0 | 0 (0.0) 0 | 0 (0.0) 0 | 0 (0.0) 0 |
| Number of subjects with at least one MAAE | 0 (0.0) 0 | 0 (0.0) 0 | 0 (0.0) 0 | 0 (0.0) 0 |
| Severity |  |  |  |  |
| Mild | 31 (19.9)<br>39 | 6 (11.5) 9 | 28 (17.9)<br>38 | 7 (13.5) 7 |
| Moderate | 4 (2.6) 5 | 1 (1.9) 1 | 0 (0.0) 0 | 1 (1.9) 1 |
| Severe | 0 (0.0) 0 | 0 (0.0) 0 | 0 (0.0) 0 | 0 (0.0) 0 |
| Life Threatening | 0 (0.0) 0 | 0 (0.0) 0 | 0 (0.0) 0 | 0 (0.0) 0 |
| Causality |  |  |  |  |
| Very likely / Certain | 19 (12.2)<br>22 | 2 (3.8) 4 | 8 (5.1) 13 | 2 (3.8) 2 |
| Probable | 2 (1.3) 2 | 0 (0.0) 0 | 7 (13.5) 9 | 2 (3.8) 2 |
| Possible | 16 (10.3)<br>19 | 5 (9.6) 6 | 15 (9.6)<br>16 | 4 (7.7) 4 |
| Unlikely | 1 (0.6) 1 | 0 (0.0) 0 | 0 (0.0) 0 | 0 (0.0) 0 |
| Unrelated | 0 (0.0) 0 | 0 (0.0) 0 | 0 (0.0) 0 | 0 (0.0) 0 |
| Unclassifiable | 0 (0.0) 0 | 0 (0.0) 0 | 0 (0.0) 0 | 0 (0.0) 0 |
| Related (Certain/Probable/Possible) | 32 (20.5)<br>43 | 7 (13.5)<br>10 | 28 (17.9)<br>38 | 8 (15.4)<br>8 |
| Unrelated (Unlikely/Unrelated/Unclassifiable) | 1 (0.6) 1 | 0 (0.0) 0 | 0 (0.0) 0 | 0 (0.0) 0 |
| Number of subjects discontinued due to AE | 0 (0.0) 0 | 0 (0.0) 0 | 0 (0.0) 0 | 0 (0.0) 0 |

### Immunogenicity Findings

Assessment of the immune response as per the primary and secondary end-points of the clinical study showed that superior immune responses were elicited in the CORBEVAX™ vaccinated cohort in comparison to the placebo cohort in both COVISHIELD™ and COVAXIN primary series recipients. The lower bound of the 95% confidence interval for the ratio of the GMT’s of nAb titers, GMC’s of Anti-RBD IgG and corresponding GMFR ratios were >1.0 in CORBEVAX™ vaccinated cohort in comparison with the placebo which demonstrates a clear statistical superiority. The percentage of subjects that showed ≥ 2-fold rise in nAb titers as well as Anti-RBD IgG concentrations also was significantly higher in CORBEVAX™ vaccinated cohort compared to the Placebo cohort (table 5 and table 6).

**Table 5:**
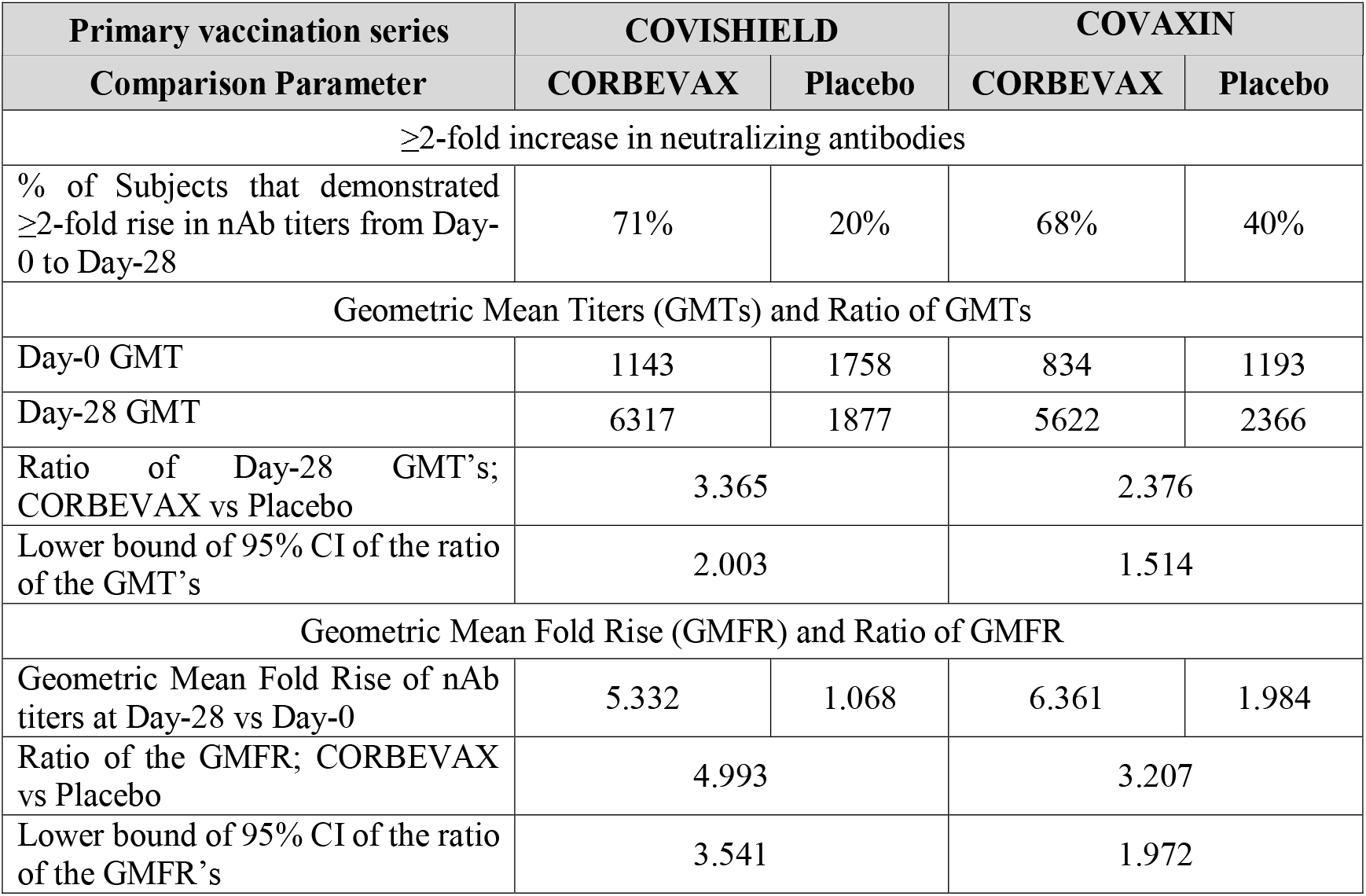
nAb titers at day 28 after booster dose with CORBEVAX vaccine vs Placebo.

**Table 6:**
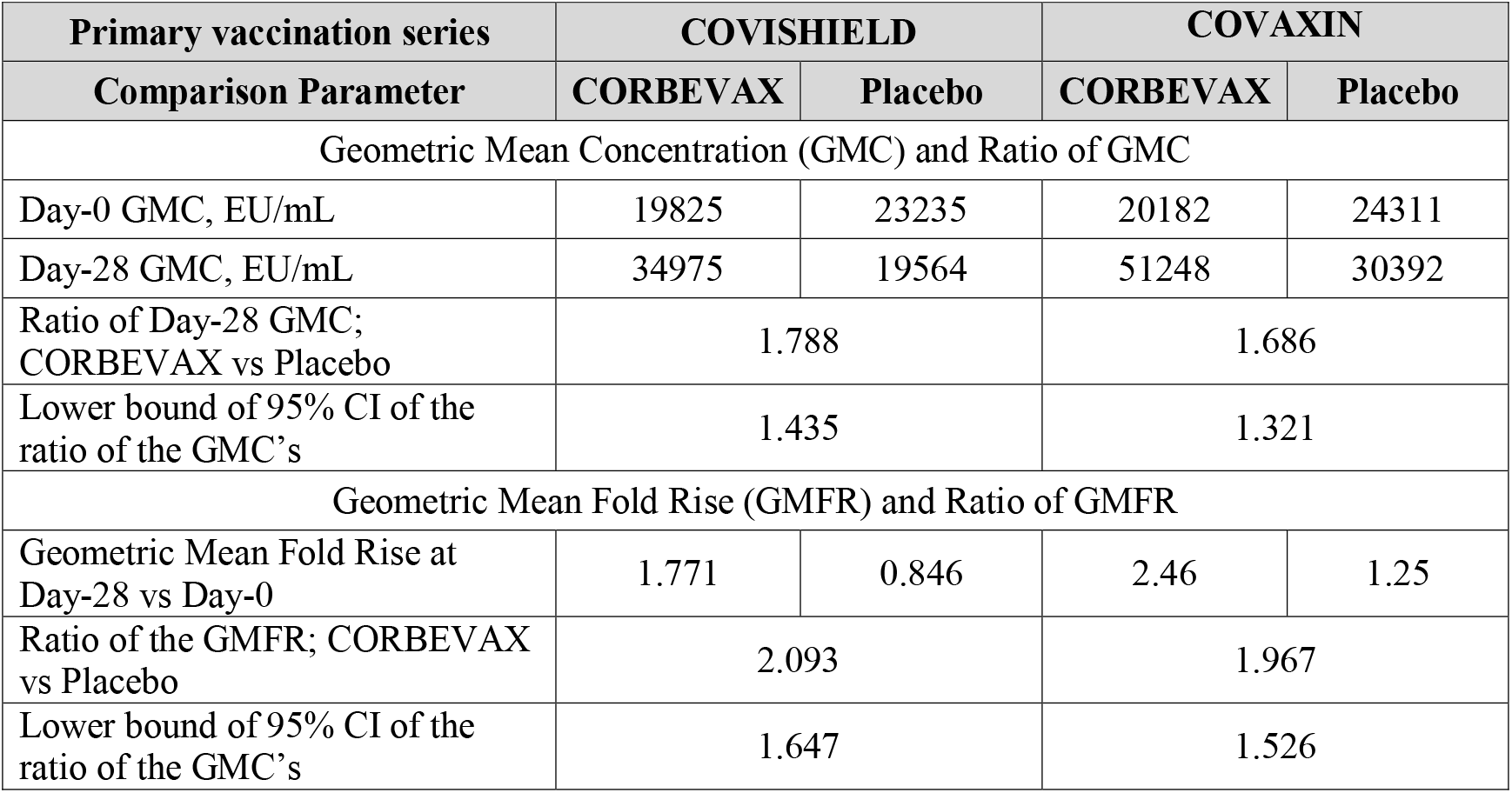
anti-RBD IgG concentrations at day 28 after booster dose with CORBEVAX vaccine vs Placebo.

As an exploratory end point, nAb titers against the omicron variant of concern were measured at baseline and day 28 after booster dose with CORBEVAX™ vaccine in both COVISHIELD™ and COVAXIN primed subjects. Significant increase was observed in terms of nAb GMT’s and percentage of subjects responded against the omicron variant of concern in CORBEVAX™ vaccinated subjects from COVISHIELD™ (day 0: 43% Vs day 28: 91%) and COVAXIN (day 0: 33% Vs day 28: 75%) recipients (table 7).

**Table 7:** Summary Omicron PNA titers pre & post Corbevax booster dose given to Covishield and Covaxin recipients.

| Assessment Parameter | Covishield | Covaxin |
| --- | --- | --- |
| Day-0 GMT | 14.9 | 11.2 |
| % Responders at Day-0 | 43% | 33% |
| Day-28 GMT | 78.8 | 45.7 |
| % Responders at Day-28 | 91% | 75% |

Cellular immune responses were assessed in a subset of subjects. The total Interferon-gamma producing PBMCs were quantified as Spot Forming Units (SFU’s) per million PBMCs using ELISPOT method. Secreted levels of IFN-gamma and IL-4 was also measured using TruCulture® assay at day 0 and day 28 after booster dose. Booster immunization with CORBEVAX™ enhanced IFN-gamma secretion (Th1 response) in both COVISHIELD™ (26.5 SFU to 79 SFU) and COVAXIN primed subjects (35.5 SFU to 219.6 SFU) as measured by EliSPOT method (table 8) while IL-4 levels (Th2 response) were comparable at baseline (day 0) and day 28 samples (table 9).

**Table 8:** IFN-gamma secretion by stimulated PBMCs at pre & post CORBEVAX booster administration-EliSPOT.

| Assessment Parameter | Covishield | Covaxin |
| --- | --- | --- |
| Average IFN-gamma secreting SFU's per million PBMC's pre-booster dose | 26.5 | 35.5 |
| Average IFN-gamma secreting SFU's per million PBMC's post-booster dose | 79 | 219.6 |

**Table 9:** Secreted cytokine data for pre & post CORBEVAX booster dose administration.

|  | Day-0<br>Null | Day-0<br>Activated | Day-28<br>Null | Day-28<br>Activated |
| --- | --- | --- | --- | --- |
| **Covishield Recipients** |  |  |  |  |
| IFN-gamma GMC; pg/mL | 2.1 | 81.6 | 1.1 | **311.5** |
| IL-4 GMC; pg/mL | 0.98 | 0.98 | 1.6 | 3.1 |
| **Covaxin Recipients** |  |  |  |  |
| IFN-gamma GMC; pg/mL | 1.97 | 82.86 | 1.66 | **127.2** |
| IL-4 GMC; pg/mL | 0.98 | 1.84 | 1.14 | 1.92 |

## DISCUSSION

In this study we report that CORBEVAX™ vaccine administered as a heterologous booster dose was safe and tolerable with no reported severe or serious adverse events. Most of the adverse events were mild in nature and related to vaccine. The safety profile is comparable to the primary vaccination series with no additional safety concerns with booster dose and safety profile observed was in line with other COVID-19 protein subunit vaccines administered as heterologous booster dose^14^.

In a previous study, it was found that ChAdOx1 did not provide protection against symptomatic disease caused by the Omicron variant after six months from administration of the second dose^2^. In a cross sectional, longitudinal study from India, it was shown that anti-spike protein antibody titers were reduced by 56% at six months after second dose of COVISHIELD™ or COVAXIN vaccines^15^. In the present study, we report that administration of heterologous booster dose with CORBEVAX™ vaccine at least six months after second dose vaccination with either COVISHIELD™ or COVAXIN significantly improved humoral and cellular immune responses and elicited superior immunogenicity compared to the placebo group. The neutralizing antibody titers were increased at least 2 fold in 71% and 68% of subjects by day28 after heterologous booster vaccination with CORBEVAX™ in COVISHIELD™ or COVAXIN primed subjects respectively. Most importantly, CORBEVAX™ booster dose has improved neutralizing antibody titers in 91% and 75% of COVISHIELD™ or COVAXIN primed subjects, respectively, against Omicron variant. These data and the data from earlier studies highlight that heterologous booster is effective in inducing neutralizing antibodies against SARS-Cov-2, including variants of concern. Major limitation of this study is that we only report boosting of immune responses at day 28 after heterologous booster dose. It is important to understand if the immune responses persist over longer duration. To address this limitation, the study subjects are being followed up for six months to evaluate long-term safety and immunogenicity. Overall, CORBEVAX™ vaccine as heterologous booster dose administered in Covishiled or COVAXIN recipients as primary series is safe, well tolerated and can induce effective humoral and cellular immune responses.

## Data Availability

Study data presented in the manuscript can be made available upon request and addressed to the corresponding author Dr. Subhash Thuluva at his.

